# Clinical sarcopenia identification: Justification for increased sensitivity in SARC-F scores for probable sarcopenia

**DOI:** 10.1101/2023.10.31.23297840

**Authors:** David Propst, Tim Dornemann, Lauren Biscardi

## Abstract

**Background:** The European Working Group on Sarcopenia in Older People (EWGSOP2) recommends the use of the 5-item SARC-F questionnaire by clinicians to screen for probable sarcopenia. The recommended threshold of ≥4 has low sensitivity and high specificity in identifying probable sarcopenia. While this high threshold is capable of excluding clients without probable sarcopenia, difficulty lies in using this screening tool to identify clients with low muscle strength.

**Methods:** 204 community-dwelling older adults (117 male, 87 female) above the age of 65 were screened at their physician visits using the SARC-F. Probable sarcopenia was diagnosed using gender-specific grip strength criteria defined by the EWGSOP2 (≤ 27 kg for men, ≤ 16 kg for women). A receiver operating characteristic (ROC) curve was used to determine a SARC-F threshold that optimized the tradeoff between sensitivity and specificity for diagnosis of probable sarcopenia.

**Results:** Probable sarcopenia was present in 11.8% of participants. There were no differences in age (73.9 ± 6.2 years) or BMI (29.5 ± 5.8 kg/m2) between genders. Males had greater grip strength (36.3 ± 8.1; 22.4 ± 5.5 kg) and lower SARC-F scores (0.92 ± 1.65, 1.88 ± 2.31) than females. The ROC curve identified a SARC-F score of ≥2 as an optimal cutoff between sensitivity and specificity (AUC = 0.77, 95% CI: 0.67 – 0.88, p < .05). Accuracy (0.77), false positive rate (0.22), positive predictive value (0.31), and negative predictive value (0.96) were also calculated.

**Conclusions:** A SARC-F threshold of ≥2 is recommended as an optimal tradeoff between sensitivity and specificity when identifying community-dwelling older adults with probable sarcopenia. This is lower than the currently accepted recommendation of ≥4. Our findings promote the recommended early detection and treatment by medical professionals by the EWGSOP2 by improving the ability to identify individuals with low muscle strength with this screening procedure.

## Introduction

Sarcopenia has been defined as a progressive loss of muscle mass and strength that adversely affects mobility, function, fall risk, and mortality in older adults (Beaudart et al., 2017; Papadopoulou, 2020; Rubbieri et al., 2014). Age-related muscle loss and strength loss can begin as early as 30 and accelerate after ages 50 and 60 (Hunt et al., 2014: Jackson et al., 2018; Rubbieri et al., 2014). The severity of muscle mass and strength in sarcopenia has been shown to be associated with decreased ability to complete activities of daily living (ADL), lower quality of life, and substantially higher healthcare costs (Cruz-Jentoft et al., 2019; Jackson et al., 2018).

In 2018, the second European Working Group on Sarcopenia in Older People (EWGSOP2) defined a multifactorial approach to identifying sarcopenia by finding, assessing, confirming, and testing the severity (Cruz-Jentoft et al., 2019). This model initially screens for sarcopenia through the use of strength, assistance with walking, rising from a chair, climbing stairs, and falls through use of a clinical symptom index (SARC-F) questionnaire or by clinical suspicion (Cruz-Jentoft et al., 2019; Malstrom et al., 2016). Individuals that are identified to potentially have sarcopenia through screening undergo a muscular strength test. If strength levels meet the criteria for sarcopenia, muscle quality testing is conducted to confirm the diagnosis (Cruz-Jentoft et al., 2019; Rubbieri et al., 2014). Then the severity of sarcopenia is determined utilizing a physical performance test (Cruz-Jentoft et al., 2019; Rubbieri et al., 2014).

The EWGSOP2 algorithm for screening and diagnosing sarcopenia includes a diagnosis of “probable sarcopenia” for use in clinical medicine. Once the SARC-F identifies possible cases then muscle strength is assessed. If muscle strength, either from grip test or chair stand test is low, then “probable sarcopenia” is diagnosed and assessment of causes may begin (Cruz-Jentoft et al., 2019).

Recognizing the importance of the diagnosis and treatment of sarcopenia in those over 65 and the impact on aging and disability, our clinic began screening all patients 65 and older at their yearly physical for probable sarcopenia. With gathered data we are looking for the answer to two questions. First, what is the prevalence of probable sarcopenia in our patient population and second, is screening for probable sarcopenia possible with use of the SARC-F screen alone?

## Methods

Two hundred and four community-dwelling older adults above the age of 65 were screened at their regularly scheduled physician visits. Participants completed a SARC-F questionnaire and a grip strength assessment. This study was approved by the university ethics committee and therefore in accordance with the ethical standards laid down in the Declaration of Helsinki and its later amendments.

### SARC-F

The SARC-F is a five question self-report survey developed by Malstrom et al. (2016) to detect clinical symptoms of sarcopenia. The SARC-F questions ask patients to report difficulties with strength, assistance walking, rising from a chair, climbing stairs, and falls. The first four items are scored as 0 (no difficulty), 1 (some difficulty), or 2 (a lot of difficulty). Number of falls in the past year is rated as 0 (no falls), 1 (between 1-3 falls), or 2 (4 or more falls). The sensitivity is low-to-moderate and the specificity is high to predict low muscle strength when a cutoff value of ≥4 is used.

### Grip Strength

Diagnosis of probable sarcopenia was assessed using the gender-specific recommended cutoff values for grip strength by the EWGSOP2 (Cruz-Jentoft et al., 2019; Dodds, 2014). These values are less than 27 kg for men and less than 16 kg for women (Dodds, 2014). Grip strength was measured using a hand-held dynamometer on the dominant hand (Sutekus Digital).

#### Statistical Analysis

Values are presented as mean and standard deviation. Normality was tested using Kolmogorov-Smirnov test and visual histograms. Comparisons between genders used between-subjects analysis. For normally distributed data, independent t-tests were used. For non-parametric data, Mann-Whitney U tests were used.

A receiver operating characteristic (ROC) curve was used to determine a threshold (SARC-F score) that optimized the tradeoff between sensitivity and specificity for diagnosing probable sarcopenia. The area under the curve (AUC) was calculated to present the ability of the SARC-F score to discriminate between probable sarcopenic and non-sarcopenic individuals. An AUC of 1.0 indicates perfect discrimination capability, 0.5 indicates discrimination capability equal to that of chance, and 0.0 indicates all subjects are incorrectly classified.

Sensitivity, specificity, positive predictive value, negative predictive value, and false positive rate were calculated for SARC-F threshold scores. Sensitivity was calculated as the number of participants diagnosed with probable sarcopenia that were correctly identified by the SARC-F screening. Specificity was calculated as the number of participants not diagnosed with probable sarcopenia that correctly screened negative with the SARC-F. Positive predictive value was calculated as the number of participants diagnosed with probable sarcopenia that screened positive with the SARC-F. Negative predictive value was calculated as the number of participants without probable sarcopenia that screened negative with the SARC-F. The false positive rate was calculated as the ratio of the number of participants screened positive by the SARC-F without probable sarcopenia to the number of participants who were not diagnosed with probable sarcopenia. Accuracy was also calculated at each SARC-F threshold. Accuracy was calculated as the proportion of correctly classified patients (both true positives and true negatives). Alpha was set at .05. Statistical analysis was performed using R Studio.

## Results

Participant characteristics are presented in Table 1. Probable sarcopenia was present in 11.8% of participants.

**Table 1.** Characteristics of study participants.

|  | Overall (N = 204) | Female (n = 87) | Male (n = 117) |
| --- | --- | --- | --- |
| Age (yrs) | 73.9 ± 6.2 | 73.8 ± 5.9 | 73.9 ± 6.4 |
| BMI (kg/m<sup>2</sup>) | 29.5 ± 5.8 | 30.2 ± 6.8 | 29.0 ± 4.8 |
| Grip strength (kg) | 30.4 ± 9.9 | 22.4 ± 5.5 | 36.3 ± 8.1* |
| SARC-F score | 1.33 ± 2.01 | 1.88 ± 2.31 | 0.92 ± 1.65* |

\* denotes significant difference from females, $p < .001$

The SARC-F cutoff score of ≥2 was identified by the ROC curve as the best tradeoff between sensitivity and specificity (Figure 1). The AUC is considered acceptable (AUC = 0.77, 95% CI: 0.67 – 0.88, p < .05). Calculations for SARC-F cutoff scores are presented in Table 2. A post hoc power analysis for the ROC curve revealed statistical power was 99.5%.

**Figure 1.**
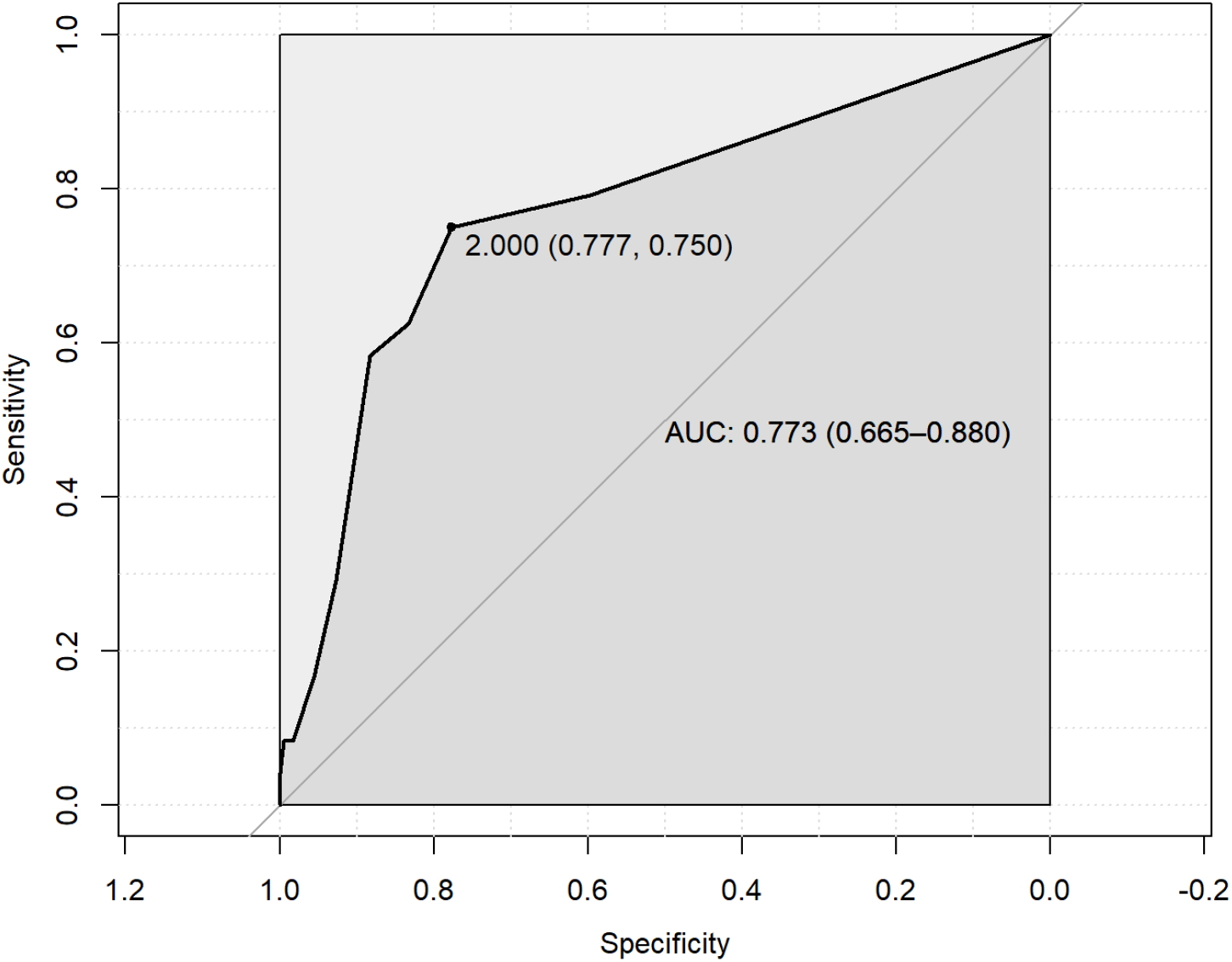
ROC curve for the SARC-F

**Table 2.** Criterion values and calculations at SARC-F thresholds.

| SARC-F<br>Cutoff Values | Sensitivity<br>(95% CI) | Specificity<br>(95% CI) | FPR | PPV | NPV | Accuracy |
| --- | --- | --- | --- | --- | --- | --- |
| 0 | 1.0000<br>(1.0000 – 1.0000) | 0.0000<br>(0.0000 – 0.0000) | 1.0000 | 0.1182 | – | 0.1182 |
| 1 | 0.7917<br>(0.6250 – 0.9583) | 0.5978<br>(0.5251 – 0.6704) | 0.4022 | 0.2088 | 0.9954 | 0.6207 |
| 2 | 0.7500<br>(0.5833 – 0.9167) | 0.7765<br>(0.7151 – 0.8324) | 0.2235 | 0.3103 | 0.9586 | 0.7734 |
| 3 | 0.6250<br>(0.4167 – 0.8333) | 0.8324<br>(0.7765 – 0.8827) | 0.1676 | 0.3333 | 0.9430 | 0.8079 |
| 4 | 0.5833<br>(0.3750 – 0.7500) | 0.8827<br>(0.8324 – 0.9274) | 0.1173 | 0.4000 | 0.9405 | 0.8473 |
| 5 | 0.2917<br>(0.1250 – 0.4583) | 0.9274<br>(0.8883 – 0.9609) | 0.0726 | 0.3500 | 0.9071 | 0.8522 |
| 6 | 0.1667<br>(0.0416 – 0.3333) | 0.9553<br>(0.9218 – 0.9832) | 0.0447 | 0.3333 | 0.8953 | 0.8621 |
| 7 | 0.0833<br>(0.0000 – 0.2083) | 0.9832<br>(0.9609 – 1.0000) | 0.0168 | 0.4000 | 0.8889 | 0.8768 |
| 8 | 0.0833<br>(0.0000 – 0.2083) | 0.9944<br>(0.9832 – 1.0000) | 0.0056 | 0.6667 | 0.8900 | 0.8867 |

Note: FPR = false positive rate; PPV = positive predictive value; NPV = negative predictive value

## Discussion

The prevalence of sarcopenia in our community-dwelling sample of older adults aligns with findings from previous studies (Papadopoulou, 2020). Average BMI indicated this sample was overweight and nearing obese. A comparison of descriptive variables between genders indicated a higher SARC-F score and lower grip strength in females, which is expected. The average grip strength for males and females in this sample both fell above the cutoff criteria for probable sarcopenia. Males reported fewer SARC-F symptoms than females. On average, both males and females were below the recommended SARC-F cutoff of 2. This is expected, due to the low prevalence of sarcopenia in the general population.

The EWGSOP2 recommends the use of the SARC-F to screen for probable sarcopenia (Cruz-Jentoft et al., 2019). This screening tool is easily implemented in the clinical setting. Prior studies rely on SARC-F cutoff of ≥4 to diagnose probable sarcopenia, which results in a high specificity but low to moderate sensitivity (Bahat et al., 2018; Cruz-Jentoft et al., 2019; Malmstrom et al., 2016). In our sample, a cutoff score of ≥4 produced moderate sensitivity (58.3%) and high specificity (88.3%) in agreement with these prior findings. As Sacar et al. (2021) noted, the SARC-F with a threshold of ≥4 is generally good at identifying people without sarcopenia (Sacar et al., 2021). Our results support this contention. However, the EWGSOP2 updates to sarcopenia diagnosis prioritize early detection and treatment for medical professionals (Cruz-Jentoft et al., 2019). A more appropriate way to meet these recommendations is to use a cutoff that better identifies individuals with probable sarcopenia.

The results of our ROC curve show that a lower cutoff value for detecting probable sarcopenia optimizes sensitivity and specificity. In agreement with the findings of Sacar et al. (2021) our ROC curve identified an optimal SARC-F cutoff of ≥2. Increasing the threshold reduces the ability of the SARC-F to detect individuals with low muscle strength and increases its ability to correctly exclude individuals without low muscle strength. To more effectively identify individuals with low muscle strength through screening, it may be better to retain sensitivity at the expense of specificity.

Recently, Dodds et al. (2020), Du et al. (2023), and Sacar et al. (2021) have argued that any positive response to the SARC-F (a threshold of ≥1) should be used to optimize SARC-F screening results. This is due to the rationale that a lower cutoff (≥1) is better to use for finding probable sarcopenia, while a higher cutoff (≥4) is better to use for excluding sarcopenia (Sacar et al., 2021). An examination of our findings in Table 2 supports this contention. As the SARC-F threshold increases, specificity increases and sensitivity decreases. These findings indicate that as the SARC-F threshold is raised, the ability of the test to correctly identify a participant with probable sarcopenia is reduced while the ability of the test to correctly identify an individual without probable sarcopenia is increased.

When it comes to the screening and diagnosis of sarcopenia, the error of a false negative (the SARC-F not identifying an individual as probable sarcopenic, when the individual is probable sarcopenic) is a more impactful error than a false positive error (the SARC-F identifying an individual as sarcopenic, when the individual is not sarcopenic). Resistance training interventions are a non-pharmacological approach successful at mitigating muscle loss and improving muscle mass, strength, and function in sarcopenic individuals (Hunt et al., 2014; Law et al., 2016; Phu et al., 2015). Beginning nonpharmacologic interventions to improve muscle strength, quality, and function, such as resistance training, can provide health benefits even to those without low muscle strength and function (Fragala et al., 2019; Hart & Buck, 2019). However, if a diagnosis is missed and the opportunity for intervention is delayed, further loss of muscle strength and function can develop. With sarcopenia screening and identification, higher sensitivity values are more desirable to reduce the occurrence of false negatives.

In conclusion, our findings indicate that screening for probable sarcopenia in community-dwelling older adults is possible with the use of the SARC-F alone. A threshold of ≥2 optimizes sensitivity and specificity, while a threshold of ≥1 prioritizes early detection. Our results, combined with earlier findings, emphasize that the clinician can confidently use the SARC-F as a simple, quick, and efficient screening strategy during routine physician visits to identify and begin treatment of probable sarcopenia without the need for time-consuming or invasive assessments. The use of the SARC-F with a lower threshold aligns the implementation of the SARC-F as a screening tool with the EWGSOP2’s recommendations for early detection and treatment of sarcopenia.

## Data Availability

All data produced in the present study are available upon reasonable request to the authors.

## Notes

### Competing Interest Statement

The authors have declared no competing interest.

### Funding Statement

This study did not receive any funding.

### Author Declarations

Barton College Institutional Review Board. The proposal, as submitted, has been deemed to meet the basic guidelines and regulations of the federal mandates and the Barton College Institutional Review Board policies.

### Summary of Updates

Correction of the displayed abstract. Correction of corresponding author details.

